# Navigating hospitals safely through the COVID-19 epidemic tide: predicting case load for adjusting bed capacity

**DOI:** 10.1101/2020.07.02.20143206

**Authors:** Tjibbe Donker, Fabian Bürkin, Martin Wolkewitz, Christian Haverkamp, Dominic Christoffel, Oliver Kappert, Thorsten Hammer, Hans-Jörg Busch, Paul Biever, Johannes Kalbhenn, Hartmut Bürkle, Winfried Kern, Frederik Wenz, Hajo Grundmann

## Abstract

**Background:** The pressures exerted by the pandemic of COVID-19 pose an unprecedented demand on health care services. Hospitals become rapidly overwhelmed when patients requiring life-saving support outpace available capacities. We here describe methods used by a university hospital to forecast caseloads and time to peak incidence.

**Methods:** We developed a set of models to forecast incidence among the hospital catchment population and describe the COVID-19 patient hospital care-path. The first forecast utilized data from antecedent allopatric epidemics and parameterized the care path model according to expert opinion (static model). Once sufficient local data were available, trends for the time dependent effective reproduction number were fitted and the care-path was parameterized using hazards for real patient admission, referrals, and discharge (dynamic model).

**Results:** The static model, deployed before the epidemic, exaggerated the bed occupancy (general wards 116 forecasted vs 66 observed, ICU 47 forecasted vs 34 observed) and predicted the peak too late (general ward forecast April 9, observed April 8, ICU forecast April 19, observed April 8). After April 5, the dynamic model could be run daily and precision improved with increasing availability of empirical local data.

**Conclusions:** The models provided data-based guidance in the preparation and allocation of critical resources of a university hospital well in advance of the epidemic surge, despite overestimating the service demand. Overestimates should resolve when population contact pattern before and during restrictions can be taken into account, but for now they may provide an acceptable safety margin for preparing during times of uncertainty.

## Introduction

The COVID-19 pandemic poses a public health threat, which, if left unmitigated, has proven potential to rapidly overwhelm health care systems^1–3^. In particular, the demand of patients that require ventilator support becomes critical when available ICU capacities are exceeded^4^. Non-pharmaceutical interventions have therefore been implemented to ameliorate the tidal wave at the peak of the epidemic (the so-called flattening of the curve)^5–7^. While national or international measures such as border closures, social-distancing, lock-downs, and furloughs have their merit in slowing the epidemic^8,9^, they also interrupt global supply chains and may thus starve health care systems from necessary equipment^10^.

Acute care, and especially tertiary care hospitals are advised to increase their capacity (beds, personnel and equipment) well in advance to cope with the expected numbers of COVID-19 patients with severe and critical conditions^11^. Although some well-known examples show that this can be achieved by creating “new” beds in temporary, purpose-build structures^12,13^, it most often is accomplished by freeing up existing bed capacity. However, this is often at the expense of hospital beds for non-COVID-19 patients^14^, and may carry opportunity costs and a protracted burden of disease. A timely estimate of the required capacity to treat COVID-19 patients is therefore critical for the planning of sufficient hospital capacity for both COVID-19 and regular patients.

The typical course of an epidemic makes early predictions about the volume and timing of peak incidence difficult due to the lack of reliable local data at a time when forecasting and planning becomes crucial. We here present how monitoring of antecedent allopatric epidemic waves, combined with timely local estimates, and continuous monitoring between March 15 and April 28 2020 helped a university tertiary care center in South-Western Germany in preparing for the pandemic and the up-scaling of bed capacity. For hospital management this offers a forecast within defined credibility boundaries allowing for better bed planning, allocation and procurement of essential resources.

## Methods

The University Medical Center Freiburg (Universitätsklinikum Freiburg, UKF) is a 1,600 bed tertiary care center and is the largest regional hospital in South-West Germany. As an acute care hospital it draws patients from about 60% of the health districts Freiburg/Breisgau/Hochschwarzwald. As a tertiary referral and trauma center, it serves other district hospitals in the Upper-Rhine region that borders Switzerland to the South and the French Alsace, Departments Haut-Rhin and Bas-Rhin, to the West.

COVID-19 cases were defined as symptomatic individuals with RT-PCR positivity for SARS-Cov-2 ascertained at one of three accredited diagnostic laboratories. Tests were carried out at community diagnostic centers, GPs, or on admission to the UKF. Positive results were reported to the district health authorities according to German notifiable disease law, and recorded as COVID-19 on hospital admission in electronic patient records. For COVID-19 patients, dates of admission, between ward referrals, and discharge were kept in electronic patient records available through the hospital patient administration system.

Prediction of the expected bed demand at the UKF was initially constrained by the availability of valid and representative data. We therefore performed a two stage approach to model the expected incidence of COVID-19 patients among the UKF catchment population. In the first stage, we used a static incidence model based on extrapolations of antecedent allopatric epidemic waves and parameterized a hospital care path model using a panel of experts. In the second stage, we used a dynamic incidence model, informed by the number of confirmed cases for the health districts Freiburg/Breisgau/Hochschwarzwald, and parameterized the care path model using the individual electronic patient records as documented by the UKF hospital patient administration system.

### Static incidence model

To provide a forecast prior to the local epidemic surge, we analyzed aggregated data from Italy and Germany as reported by the John Hopkins University^15^, as well as sub-national data for the region Lombardy and the province of Lodi available through the website of the Italian Dipartimento della Protezione Civile^16^. We calculated the delay between the cumulative per capita incidence in each region relative to the Italian epidemic, and normalized the epidemic curves by correcting for this delay, thus overlaying and combining all curves into a single epidemic trajectory. We tested the trajectory for saturation properties and decided on a tentative epidemic peak on which we mirrored the epidemic curve, following a symmetry conjecture.

To predict the Freiburg regional epidemic, we calibrated the epidemic curve to the UKF catchment population (taking into account observed delays) by multiplying the per capita values with the catchment population size of the UKF. Calculation of catchment size was based on relative number of admissions to the UKF in comparison to all other hospitals in the state of Baden-Württemberg (11. Mio inhabitants) taken from a comprehensive dataset recording all patient admissions on an annual basis. The database was made available by the largest health insurer (Allgemeine Ortskrankenkasse Baden-Württemberg, AOK-BW).

### Dynamic incidence model

We produced local data informed estimates, using the dynamic incidence model, after cumulative case counts reached 1/1,000 in the Freiburg/Breisgau/Hochschwarzwald health districts (492,000 inhabitants). We imputed the likely date of infection for each COVID-19 case in the health district and estimated the time-varying effective reproduction number (R_T_)^17^ based on the probability distribution of the serial intervals between consecutive case generations^18^. We used an individual-based stochastic simulation to predict the local future incidence, assuming that R_T_ declined exponentially over time, fitting R_f_ (t) = a e^bt^ to the estimated R_T_. This declining function serves as a phenomenological approximation to the observed changes in R_T_ and is used to calculate the transmission parameter of the SIR model (see supplementary text S1).

### Care path model

To convert the forecasted regional incidence to bed demand, we created an agent-based model for the in-hospital care path (Figure 1), consisting of: (C) Confirmed cases, equal to the results of the above incidence models, (GW) patients admitted to general wards, and (ICU) patients admitted to intensive care units. Patients are assumed to follow one of three possible tracks through the hospital: 1) admitted and discharged to and from GW. 2) admitted to GW, moved to ICU, and then discharged from ICU, or 3) directly admitted to ICU and discharged from there. Within the model, we make no distinction between discharge and death, and removed patients as end of stay. The model thus contains 5 parameters: the distributions of the length of stay in the GW and ICU, the distribution of time from infection to hospital admission, the proportion of patients admitted to hospital, and the proportion of patients directly admitted to ICU.

**Figure 1:**
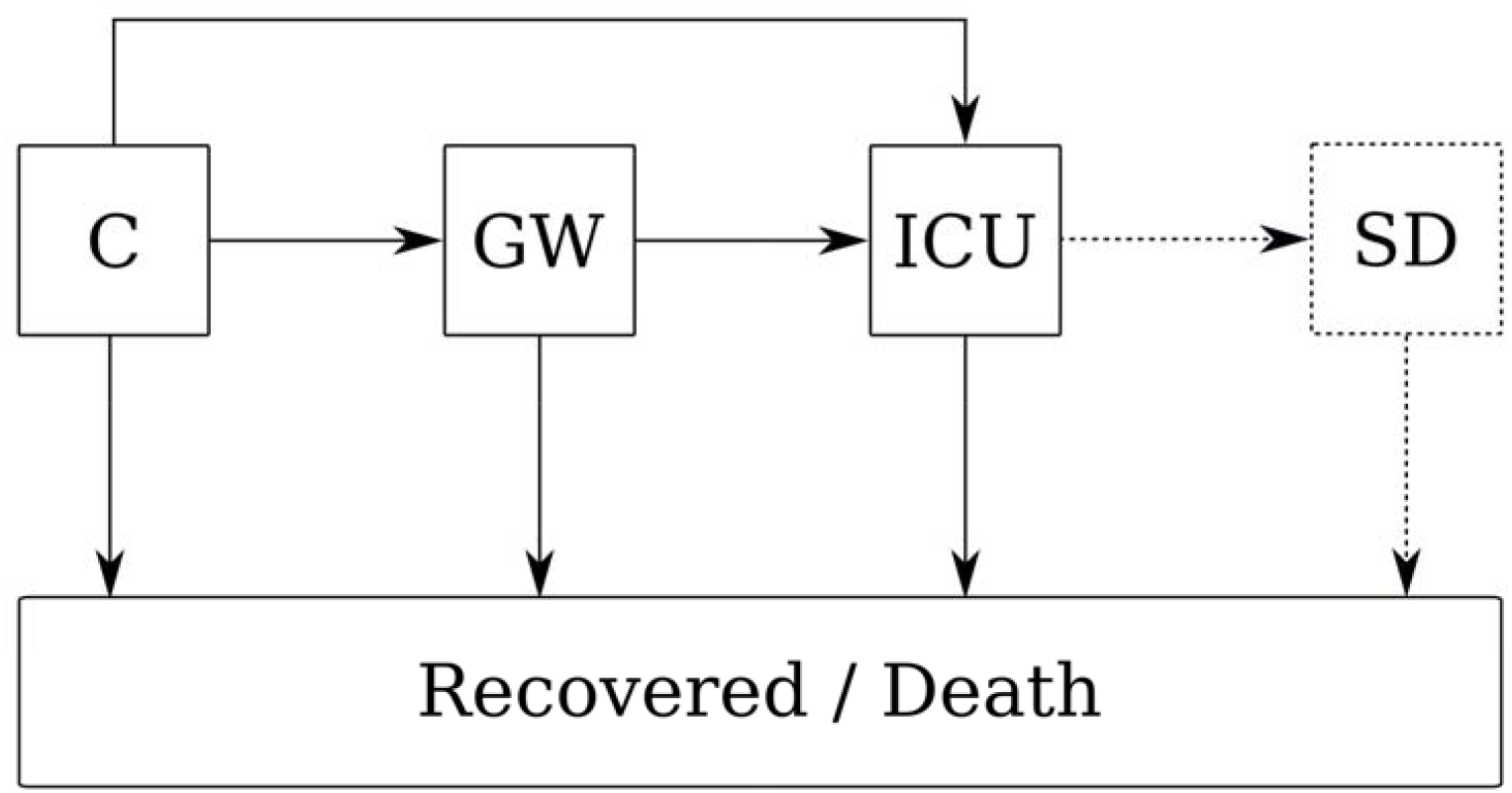
Model structure. A) The COVID-19 care path, describes how patients progress from confirmed cases in the community (C), to be admitted on general wards (GW), to intensive care units (ICU), and to step-down units (SD). Some COVID-19 patients are admitted directly to the ICU from the community. The step-down unit was only included in the agent-based model.

In the first stage, we used a consensus care-path parameterized by judgment of a panel of experts. In absence sufficient local data, we asked 4 consultant experts (3 intensivists, 1 infectious diseases specialist) to make estimates about the expected care path of COVID-19 patients during their treatment in the UKF.

Once the number of admitted COVID-19 patients had surpassed 150, we parameterized the empirical care-path using the individual electronic patient records as documented by the UKF hospital patient administration system. Based on these observations, we added a fourth compartment for those patients who left the ICU and returned to a general ward and named this a step-down unit (SD).

We analyzed the time until end of stay in each compartment split between those patients being transferred to another compartment and those ending their stay. We fit both exponential and Weibull distributions to the hazard functions of leaving each compartment. These served as the main parameters in the empirically informed care-path. Furthermore, we calculated the admission rate as the cumulative number of admissions divided by the cumulative number of confirmed cases, and similarly, we determined the proportion of patients being directly admitted to the ICU.

## Results

During the first two weeks of March 2020, the region of Lombardy in Italy and the Departments Haut-Rhin and Bas-Rhin in France saw a rapid expansion of the COVID-19 epidemic^1,19,20^. The civil protection authorities in Italy reported confirmed cases on a daily basis. We observed that cumulative case counts for Germany followed the same exponential trajectory as the province of Lodi, the region of Lombardy and Italy as a whole, albeit with some delay but similar growth rates (Figure 2A&B). The delay between Germany and Italy was estimated to be 10 days, while Lodi province was 21 days ahead of Italy. As shown in Fig. 2A&B only the trajectory for Lodi showed an obvious and sustained slowing of the growth rate prior to March 15. We chose March 7 as the saturation point for the epidemic in Lodi province, and assuming an equivalent dynamic, the peak for Germany could be projected to occur on April 7. Applying the combined trajectory (Figure 2C) to the catchment population of the UKF (290.000 people), we expected 103 incident cases on the day of the epidemic peak.

**Figure 2:**
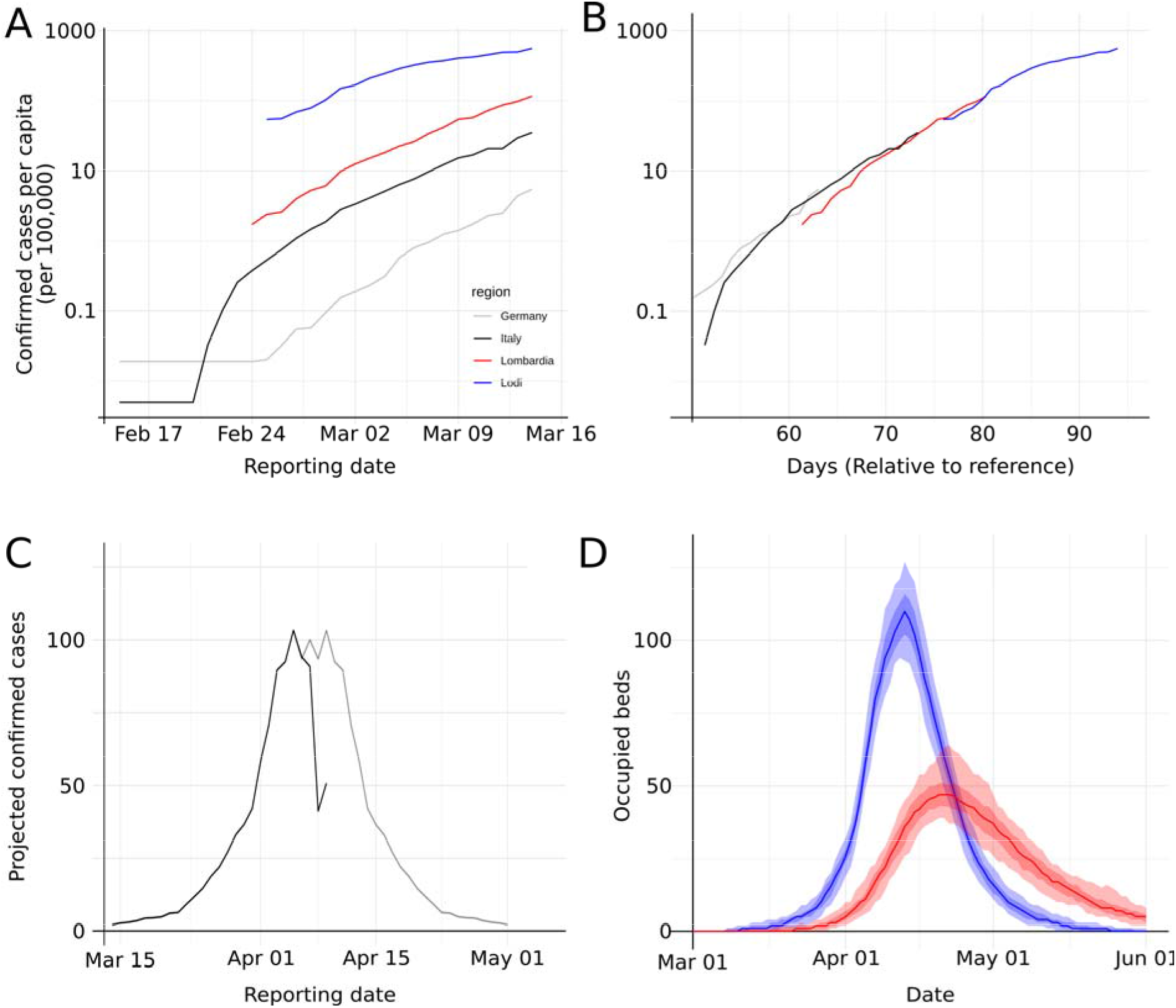
Early forecast using the static model. A) The trajectory of the number of confirmed cases in Germany, Italy, Region of Lombardy, and Province of Lodi were B) normalized and projected as a single curve by compensating for the apparent delay between locations, and C) the downward slope (grey) was predicted assuming a symmetry conjecture of the observed upward slope (black). D) Expected bed occupancy, (general wards in blue, ICU in red). Light shades 95% CI, dark shades interquartile ranges. Predictions are based on The COVID-19 care path using expert consensus. Bed demand peaked on the general wards at 116 beds on April 9 and in ICUs at 47 beds on April 19.

In order to predict the expected bed and ventilator demand we chose to describe the expected COVID-19 patient care-path on the basis of expert knowledge and opinion. The expert estimates were in general agreement on most of the care-path parameters, with a couple of exceptions (see table S2). We combined the assessments by averaging the individual parameter estimates of all 4 experts into a single consensus care-path. Combining the care-path with the results of the static incidence model, we predicted the demands for general ward and ICU beds to peak on April 9 (116 beds) and April 19 (47 beds), respectively (Figure 2D).

By April 5, the availability of locally generated data provided the opportunity to populate the care-path model with empirical local data and offer the first predictions using the dynamic incidence model. At the time, 153 patients had been admitted to the UKF, of which 55 required ventilator support on ICUs. 28 were admitted directly to an ICU, whereas 27 had a prior stay on a GW. 14 had already been transferred to the SD. Of all admitted patients, 87 were still hospitalized (GW:48, ICU:29, SD:10). Based on the estimated hazard of end of stay on the GW (0.05678 d^-1^) and the hazard of transfer to the ICU (0.0307 d^-1^), we estimated that patients spent a mean of 11.4 days on the GW (Figure 3A&D). Similarly, we estimated patients stayed on average 14.7 days on the ICU (end of stay hazard: 0.03127, transfer hazard: 0.03648; Figure 3B&E), and 40.5 days on the step down unit (Figure 3C; based on 4 discharges). The hazard estimates stabilized over time, as more data were added (Figure 3F, table S3).

**Figure 3:**
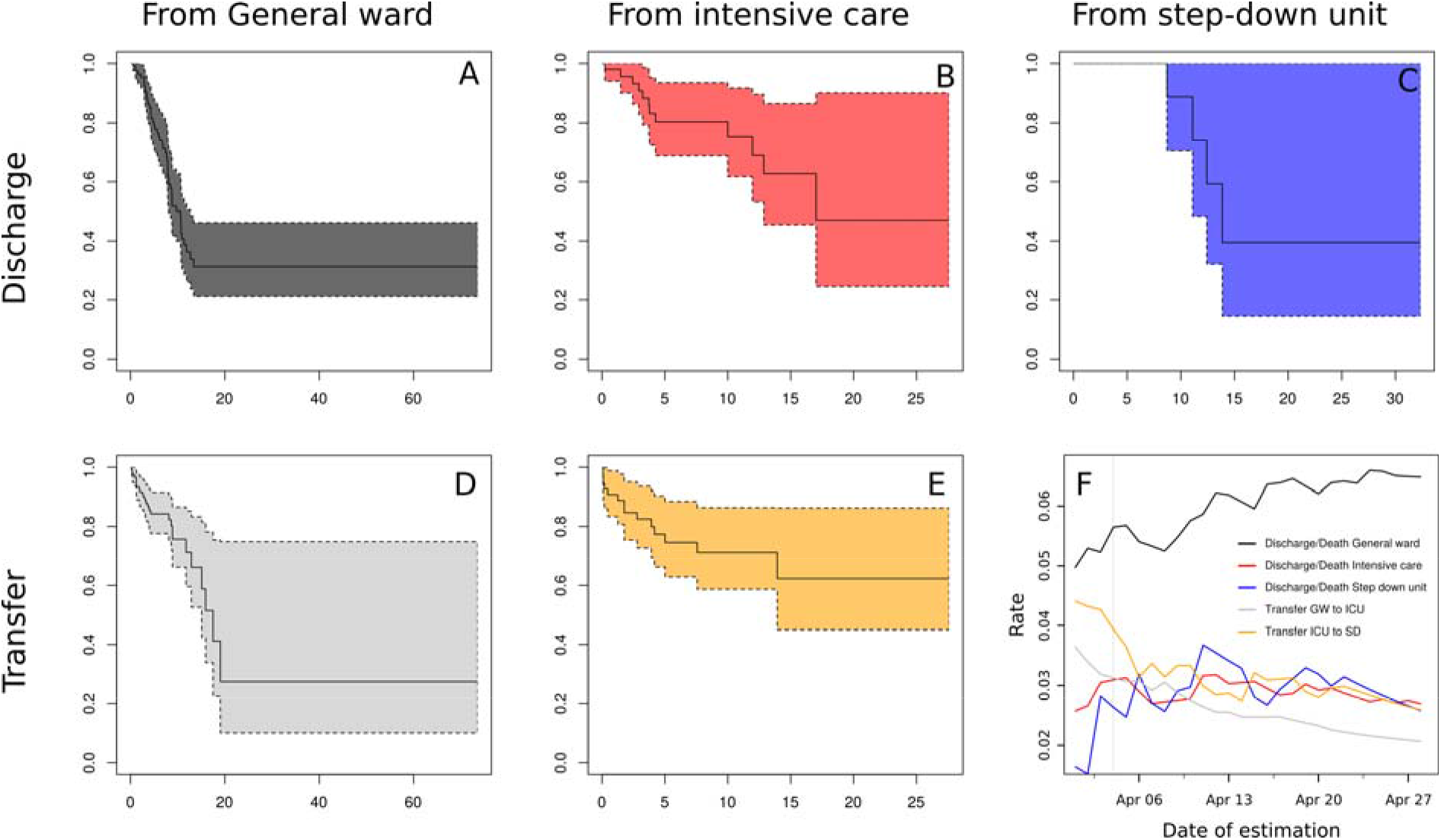
Survival analysis A-E) Kaplan-Meier estimators for the stay on the general ward (A & D), ICU (B & C), and step-down unit (C), for patients that are discharged (A,B & C) and transferred to the following ward (D&E), based on the data observed on April 5. (F) The estimated rates of discharge/death and transfers over time, based on continuously accumulating data.

On the same day, 1372 cumulative cases (Figure 4A) had been reported in the health district (2.8 per 1.000 inhabitants). Since the onset of the epidemic the time-varying reproduction number (R_T_) showed a clear decline, starting at a median of 3.5, and decreased to 1.1 (Figure 4B). The dynamic incidence model projected a peak incidence of a median number of 90 cases for April 8 (Figure 4C). Although there was considerable variation between model simulations (the lowest peak was projected at 74 cases, the highest at 1186), 75% of the realizations suggested little or no further increase with an imminent saturation of the epidemic in the near future. Dynamic incidence forecasts could be produced from March 24, albeit without empirical local care path data until April 5. While adding daily reported cases to the dynamic incidence model, iterations generated fluctuations and occasional uncertainty (see Figure S4, Movie S5). The overall trajectory stabilized after April 6. Combining these projections with the empirically informed care-path, we estimated a peak demand of 102 general ward beds (IQR = 92-121) on April 17 and 49 ICU beds (IQR = 42-58) on April 25 (Figure 4D). Observed bed occupancy peaked on April 8 for both the general wards (66 beds occupied) and the ICU (34 beds occupied). By April 14, the forecasted bed demand aligned with the observed occupancy (see figure S4, 5^th^ column), likely because the incidence model predicted the declining phase of the epidemic curve more precisely.

**Figure 4.**
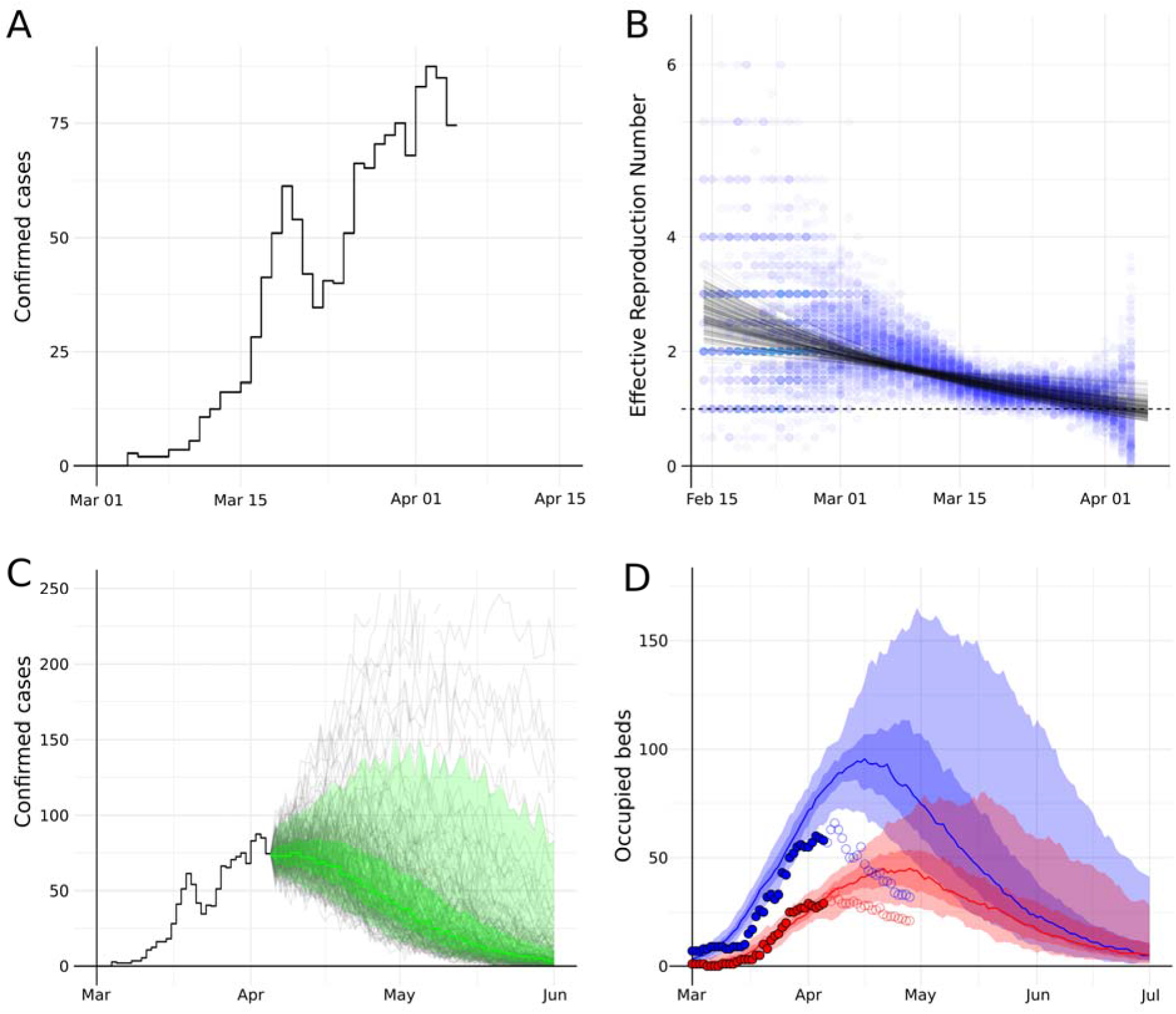
Late forecast using the dynamic model based on locally available data on April 5. A) The observed incidence of confirmed cases in the health districts Freiburg/Breisgau/Hochschwarzwald combined. B) Backward model: Estimates of the time-varying Reproduction number (blue dots) over 100 stochastic simulations, with fitted R_f_(t) trajectories (black lines). C) Forward model: Forecasted incidence of confirmed cases, grey lines show single simulation results, green line show the median, with green shade showing the interquartile range and green light shade 5%-95% of the simulation results. D) Estimated bed demand (median, IQR, 5%-95% range) for the general wards (blue) and ICU (red). Circles denote actual observed number of beds occupied (closed: past days, open: future days not know at the time of the analysis on April 5).

## Discussion

During the current COVID-19 pandemic, hospitals have reported a break-down of services when the surge of patients in need of treatment and ventilator support outpaced available capacities^21^. Early predictions about the timing and volume of maximum service demand, i.e. expected general ward and ICU bed occupancy, are therefore critical in the early stages of an epidemic. These may help guide the upscaling of a hospital’s bed capacity, redistributing personnel, and storage of crucial equipment. However, data that could provide a basis for predictions are often equivocal or insufficient in the incipient stages of an epidemic. In an attempt to decrease these uncertainties, we have utilized allopatric and locally available data for contingency planning for health care authorities and hospital management.

We took advantage of antecedent epidemic waves in neighboring countries, especially in Northern Italy where daily case counts were available at the level or provinces with population sizes comparable to our own health district. When we normalized the epidemic trajectories between nations, regions and provinces, we found the notion confirmed that uncontrolled transmission in populations with similar contact patterns^22^ is comparable during early stages of epidemics. We therefore informed our local predictions by a static model calibrated by the per capita case counts and delay of onset in Lodi, Lombardy and Italy. This gave local health care authorities and hospital management of the UKF three weeks to prepare for the expected number of COVID-19 patients at the epidemic peak, ample time to call-off elective interventions, upscale ICU capacity, reallocate staff, take stock, and order essential equipment such as, disposables, personnel protective equipment (PPE), oxygen etc. Given the size of the epidemics in Lombardy and the neighboring department in France, the results of the static model were used as the lower bound of the required capacity, as it was uncertain if regional district hospitals would be able to cope with the likely caseloads in the larger region. The entire region consists of a catchment population three times the size of the UKF’s catchment (1 Mio vs 290k), which was forecasted to result in an additional demand for 284 GW and 121 ICU beds in the surrounding hospitals (Figure S6).

The dynamic model required locally available data. After March 24, sufficient numbers of incident cases had been ascertained and moved through the UKF, allowing forecasts to be updated on a daily basis. This real-time tracking provided us with the potential to adjust hospital planning. When forecasts became ambiguous between April 1 and April 6, the decision was taken to closely monitor the development at weekly intervals, since the planning based on the static model set aside enough ICU beds available for COVID-19 patients and regional district hospitals were still coping. In hindsight, our model predicted the peak to be higher and later than the one observed.

The implicit continuity assumption of some of the parameters made our prediction models vulnerable to unforeseen changes in transmission dynamics, such as the introduction of NPIs. For Freiburg and surroundings, restrictions took effect on March 20, and included closures of schools, shops and restaurants, prohibition of large gatherings, and an obligation for social distancing. Our fitted trajectory to the estimated time-varying reproduction number (R_T_) may therefore initially have been too high, as pre-intervention R_T_ estimates still carried too much weight. Additionally, random mixing assumed in the standard SIR model overestimates epidemic growth, since extant contact pattern are generally assortative. Furthermore, the incidence model assumes homogeneity in the patient population, i.e. each infected individual having the same probability of being admitted to hospital. In reality, risks are strongly associated with age^23–25^ which skews hospitalization rates depending on demographic composition. We therefore suggest that improved prediction models should account for age stratified contact patterns, age structure of hospital catchment populations, and the effect of NPIs.

Local data based short-term forecasts of hospital admissions are vital to epidemic planning of hospitals, as the onset of epidemics may vary greatly between different geographical regions. With that, local bed demand may peak at different times. The lack of local data in the early phase of an epidemic is challenging in this respect. We solved this issue by forecasting in two stages, with an early “crude” estimate based on observed COVID-19 outbreaks abroad, and a later, continuously adjusted nowcasting and forecasting by incorporating locally ascertained data. This way, we were able to navigate hospital capacities by setting weekly targets, while adjusting the elective admission and discharge policies according to the most current epidemic situation.

## Data Availability

All data needed to perform the analysis is provided, with the exception of the raw patient care path data, for privacy reasons.

https://github.com/tjibbed/UKFprojection

## Authors’ contributions

Study conception: TD, FB, HG. Data collection, CH, DC, OK, TH, HJB, PB, JK, HB, WK. Data interpretation: TD, FB, MW, TH, HJB, PB, JK, HB, WK, FW, HG, Writing of manuscript: TD, HG. Critical revision of the manuscript: All authors.

## Conflict of interest

We declare no competing interests.

## Role of funding source

This study had no funding source. The corresponding author had full access to all the data in the study and had final responsibility for the decision to submit for publication.

## Notes

### Competing Interest Statement

The authors have declared no competing interest.

## References

1. Guzzetta G, Poletti P, Ajelli M, et al. Potential short-term outcome of an uncontrolled COVID-19 epidemic in Lombardy, Italy, February to March 2020. Eurosurveillance. 2020;25(12):2000293. doi:10.2807/1560-7917.ES.2020.25.12.2000293

2. Li R, Rivers C, Tan Q, Murray MB, Toner E, Lipsitch M. The demand for inpatient and ICU beds for COVID- 19 in the US: lessons from Chinese cities. medRxiv. Published online March 16, 2020:2020.03.09.20033241. doi:10.1101/2020.03.09.20033241

3. WHO. Critical preparedness, readiness and response actions for COVID-19. Published March 19, 2020. Accessed May 26, 2020. https://apps.who.int/iris/bitstream/handle/10665/331494/WHO-2019-nCoV-Community_Actions-2020.2-eng.pdf

4. Grasselli G, Pesenti A, Cecconi M. Critical Care Utilization for the COVID-19 Outbreak in Lombardy, Italy: Early Experience and Forecast During an Emergency Response. JAMA. 2020;323(16):1545–1546. doi:10.1001/jama.2020.4031

5. Cowling BJ, Ali ST, Ng TWY, et al. Impact assessment of non-pharmaceutical interventions against coronavirus disease 2019 and influenza in Hong Kong: an observational study. Lancet Public Health. 2020;5(5):e279–e288. doi:10.1016/S2468-2667(20)30090-6

6. Matrajt L, Leung T. Evaluating the Effectiveness of Social Distancing Interventions to Delay or Flatten the Epidemic Curve of Coronavirus Disease. Emerg Infect Dis. 2020;26(8). doi:10.3201/eid2608.201093

7. Ferguson N, Laydon D, Nedjati Gilani G, et al. Report 9: Impact of Non-Pharmaceutical Interventions (NPIs) to Reduce COVID19 Mortality and Healthcare Demand. Imperial College London; 2020. doi:10.25561/77482

8. Davies NG, Kucharski AJ, Eggo RM, Gimma A, CMMID COVID-19 Working Group, Edmunds WJ. The Effect of Non-Pharmaceutical Interventions on COVID-19 Cases, Deaths and Demand for Hospital Services in the UK: A Modelling Study. Infectious Diseases (except HIV/AIDS); 2020. doi:10.1101/2020.04.01.20049908

9. White ER, Hébert-Dufresne LR. State-level variation of initial COVID-19 dynamics in the United States: The role of local government interventions. medRxiv. Published online April 17, 2020:2020.04.14.20065318. doi:10.1101/2020.04.14.20065318

10. Livingston E, Desai A, Berkwits M. Sourcing Personal Protective Equipment During the COVID-19 Pandemic. JAMA. 2020;323(19):1912–1914. doi:10.1001/jama.2020.5317

11. Guan W, Ni Z, Hu Y, et al. Clinical Characteristics of Coronavirus Disease 2019 in China. N Engl J Med. 2020;382(18):1708–1720. doi:10.1056/NEJMoa2002032

12. London’s ExCel centre will treat Covid-19 patients “within days.” The Guardian. https://www.theguardian.com/world/2020/mar/24/londons-excel-centre-will-be-treating-covid-19-patients-within-days. Published March 24, 2020. Accessed May 26, 2020.

13. Inside the Javits Center: New York’s militarized, makeshift hospital. Washington Post. Accessed May 26, 2020. https://www.washingtonpost.com/national/javits-center-coronavirus-field-hospital/2020/04/04/50bdbf32-75b2-11ea-87da-77a8136c1a6d_story.html

14. Rosenbaum L. The Untold Toll — The Pandemic’s Effects on Patients without Covid-19. N Engl J Med. 2020;0(0):null. doi:10.1056/NEJMms2009984

15. CSSEGISandData. CSSEGISandData/COVID-19.; 2020. Accessed May 26, 2020. https://github.com/CSSEGISandData/COVID-19

16. Pcm-Dpc/COVID-19. Presidenza del Consiglio dei Ministri - Dipartimento della Protezione Civile; 2020. Accessed May 26, 2020. https://github.com/pcm-dpc/COVID-19

17. Wallinga J, Teunis P. Different epidemic curves for severe acute respiratory syndrome reveal similar impacts of control measures. Am J Epidemiol. 2004;160(6):509–516. doi:10.1093/aje/kwh255

18. Cereda D, Tirani M, Rovida F, et al. The early phase of the COVID-19 outbreak in Lombardy, Italy. ArXiv200309320 Q-Bio. Published online March 20, 2020. Accessed May 26, 2020. http://arxiv.org/abs/2003.09320

19. Bonnasse-Gahot L, Dénès M, Dulac-Arnold G, et al. ICUBAM: ICU Bed Availability Monitoring and analysis in the Grand Est region of France during the COVID-19 epidemic.:14.

20. Breton V, Guiguet-Auclair C, Odoul J, Peterschmitt J, Ouchchane L, Gerbaud L. Population Based Survey of the COVID-19 Outbreak in the Haut-Rhin Department from January to April 2020. Social Science Research Network; 2020. Accessed May 26, 2020. https://papers.ssrn.com/abstract=3601684

21. Phua J, Weng L, Ling L, et al. Intensive care management of coronavirus disease 2019 (COVID-19): challenges and recommendations. Lancet Respir Med. 2020;8(5):506–517. doi:10.1016/S2213-2600(20)30161-2

22. Mossong J, Hens N, Jit M, et al. Social Contacts and Mixing Patterns Relevant to the Spread of Infectious Diseases. Riley S, ed. PLoS Med. 2008;5(3):1.

23. Yang X, Yu Y, Xu J, et al. Clinical course and outcomes of critically ill patients with SARS-CoV-2 pneumonia in Wuhan, China: a single-centered, retrospective, observational study. Lancet Respir Med. 2020;8(5):475–481. doi:10.1016/S2213-2600(20)30079-5

24. Wang D, Hu B, Hu C, et al. Clinical Characteristics of 138 Hospitalized Patients With 2019 Novel Coronavirus–Infected Pneumonia in Wuhan, China. JAMA. 2020;323(11):1061–1069. doi:10.1001/jama.2020.1585

25. Garg S. Hospitalization Rates and Characteristics of Patients Hospitalized with Laboratory-Confirmed Coronavirus Disease 2019 — COVID-NET, 14 States, March 1–30, 2020. MMWR Morb Mortal Wkly Rep. 2020;69. doi:10.15585/mmwr.mm6915e3

